# Estimating recent trends in UK alcohol sales from alcohol duty revenue

**DOI:** 10.1101/2024.12.12.24318899

**Authors:** Colin Angus, Jonas Schöley

## Abstract

**Background and Aims:** The onset of the COVID-19 pandemic led to significant changes in individual-level alcohol consumption and a sharp increase in heavy drinking in the UK. More recently, high rates of inflation, the resulting ‘cost-of-living crisis’ and reforms to alcohol taxation have affected the affordability of alcohol, but little is understood about how these changes have impacted on alcohol sales and consumption. We aimed gain insights into recent trends in alcohol sales by assessing changes in alcohol duty revenue collected by the UK government since 2020.

**Design and Setting:** We used published data on UK alcohol duty revenue to model trends from 2010-2019. We forecasted these trends through to October 2024 using a novel statistical approach and compared these forecasts to observed receipts.

**Measurements:** Monthly inflation-adjusted alcohol duty receipts received by the UK Treasury in pounds sterling for beer, cider, spirits and wine.

**Findings:** During the pandemic, alcohol duty receipts fell during lockdowns and rose as restrictions were subsequently lifted. Since 2022 alcohol duty receipts have been consistently below the historical trend, with a gradually increasing deficit in wine receipts and comparable deficit in spirits receipts that began sharply in late 2022. The reforms to the alcohol duty system in August 2023 do not appear to have significantly affected these trends.

**Conclusions:** Our findings suggest that the ‘cost of living crisis’ in 2022/23 has led to a fall in alcohol sales relative to the pre-pandemic trend. The magnitude of this fall differs by beverage type, indicating that wine and spirits drinkers may have changed their behaviour more than beer and cider drinkers.

## Introduction

The harms associated with alcohol consumption place a substantial burden on society, with total societal costs estimated to exceed £27billion per year in England^1^. The onset of the COVID-19 pandemic in 2020 was associated with marked changes in alcohol consumption for many individuals, with multiple studies finding a polarisation in drinking, with heavier drinkers drinking more and moderate drinkers consuming less alcohol, or giving up entirely^2–5^. Alongside these changes, the United Kingdom (UK) has seen a sharp increase in deaths from wholly alcohol-attributable causes, rising by 32.8% between 2019 and 2022 to their highest level on record^6^.

Beyond the pandemic, inflation rates in the UK began to rise in 2021, reaching a 40-year high in October 2022, leading to a so-called ‘cost-of-living crisis’ as the costs of goods and energy rose faster than incomes^7^. Further, in August 2023, the UK Government introduced reforms to alcohol taxation, designed with the explicit goal of improving public health. Under these reforms all alcohol products are now taxed on the basis of their alcohol content, with duty rates rising with alcoholic strength in line with recommendations from the World Health Organization^8,9^. Although these reforms were designed to be broadly revenue neutral for government, at the same time as their implementation alcohol duty rates were increased by 10.1% in line with inflation, the largest single increase in alcohol taxes for over 40 years.

To date, studies on changes in alcohol consumption since the start of 2020 have used individual-level survey data, that may be subject to self-report bias, and focused on changes during COVID restrictions in the early phase of the pandemic. Little is known about how consumption may have changed in the subsequent period during the cost-of-living crisis or in response to the alcohol duty reforms.

In order to understand the impact of these diverse events on alcohol consumption and the potential implications for future rates of alcohol harms, we examined UK government data on alcohol duty revenues, comparing revenues since 2020 to the pre-pandemic trend. Duty revenues represent a strong proxy measure for the volume of alcohol sold in the UK as the majority of alcohol is taxed on the basis of its alcohol content.

## Methods

In order to detect unusual alcohol consumption patterns since 2020 we estimate the deviation between observed and expected UK alcohol duty receipts. We used monthly data on UK alcohol duty receipts published by HM Revenue and Customs for the period from January 2010 to October 2024^10^. Data is published separately for duty receipts from beer, cider, spirits and wine. All values were inflated to October 2024 prices using the Retail Prices Index, the inflation measure used to uprate duty rates annually^11^. Deviations in receipts since 2020 relative to pre-pandemic trends, by beverage type, were assessed using a methodological approach that has previously been applied to estimate cause-specific excess mortality during the pandemic^12^. The calculation of expected receipts involved a two-stage process.

First, using data from January 2010 to December 2019, we modelled the expected total monthly inflation adjusted duty revenues at time 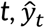, as a log-linear function of year and month via

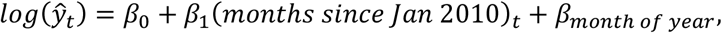

where *β*_1_ is the slope of a log-linear long-term trend and *β*_*month of year*_ is a month-specific fixed effect, to account for seasonality.

Second, we fit a compositional regression to estimate the expected proportion of total duty receipts that come from each beverage type *i* at time 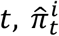, using the isometric log-ratio transformation^13^

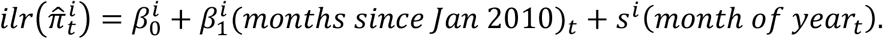

We can then estimate the expected receipts for any beverage type *i* at time *t* as

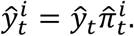

A multivariate normal distribution of errors in the predicted duty revenues, correlated across beverage types, has been estimated from the residuals of the fitting data. In order to quantify the uncertainty around our estimates we perform a residual bootstrap from this distribution.

The compositional approach was chosen to ensure coherency between expected duty revenues on both the beverage specific and the total level^14^. Duty revenues by beverage sum up to total revenues and prediction intervals, being derived from correlated errors, have good calibration on both the beverage specific level of analysis as well as the total level.

Based on the fitted models we predict expected receipts by beverage type for the months following Jan 2020. Deviations from the expectation are the given by simple difference between observed and expected duty revenues.

See the Online Supplementary Material for diagnostic plots of model fit (Figures A1 and A2). An inspection of long-term trends in duty revenue by beverage type (Figure A3) suggests the possibility that trends in wine revenues may have changed in early 2018. We therefore undertook a sensitivity analysis where we used data up to December 2017 as the training data and assessed deviation from this trend from January 2018 onwards. All analyses were performed using R statistical software. Full code to replicate the analysis can be found at https://github.com/VictimOfMaths/Experiments/blob/master/HMRCReceiptsAnalysis.R. This analysis was not pre-registered and should therefore be considered exploratory.

## Results

Figure 1 shows the monthly deviation from expected, or ‘excess’, duty revenue collected by UK Government since January 2020. This illustrates that duty receipts fell sharply in March and April 2020, when pandemic lockdown measures were first introduced, but this was offset by a corresponding increase in revenues in the summer of 2020 as restrictions were lifted. This pattern was repeated in late 2020/early 2021 when lockdowns were reintroduced, and subsequently relaxed in spring 2021. Figure 2 shows the cumulative difference between observed and expected receipts since January 2020, illustrating that the net impact of these changes in duty receipts across 2020 and 2021 was close to zero.

**Figure 1.**
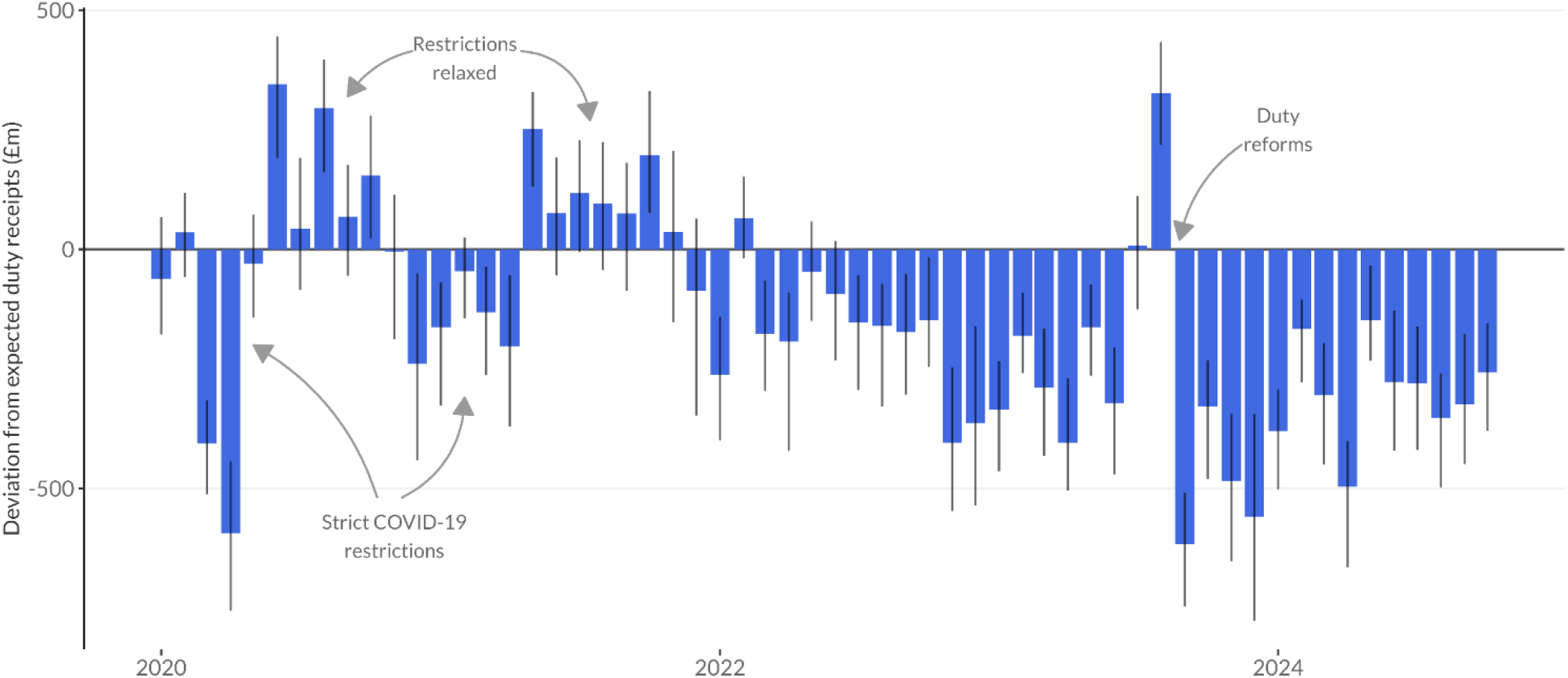
Excess monthly duty revenue in the UK since January 2020 with 95% prediction intervals.

**Figure 2.**
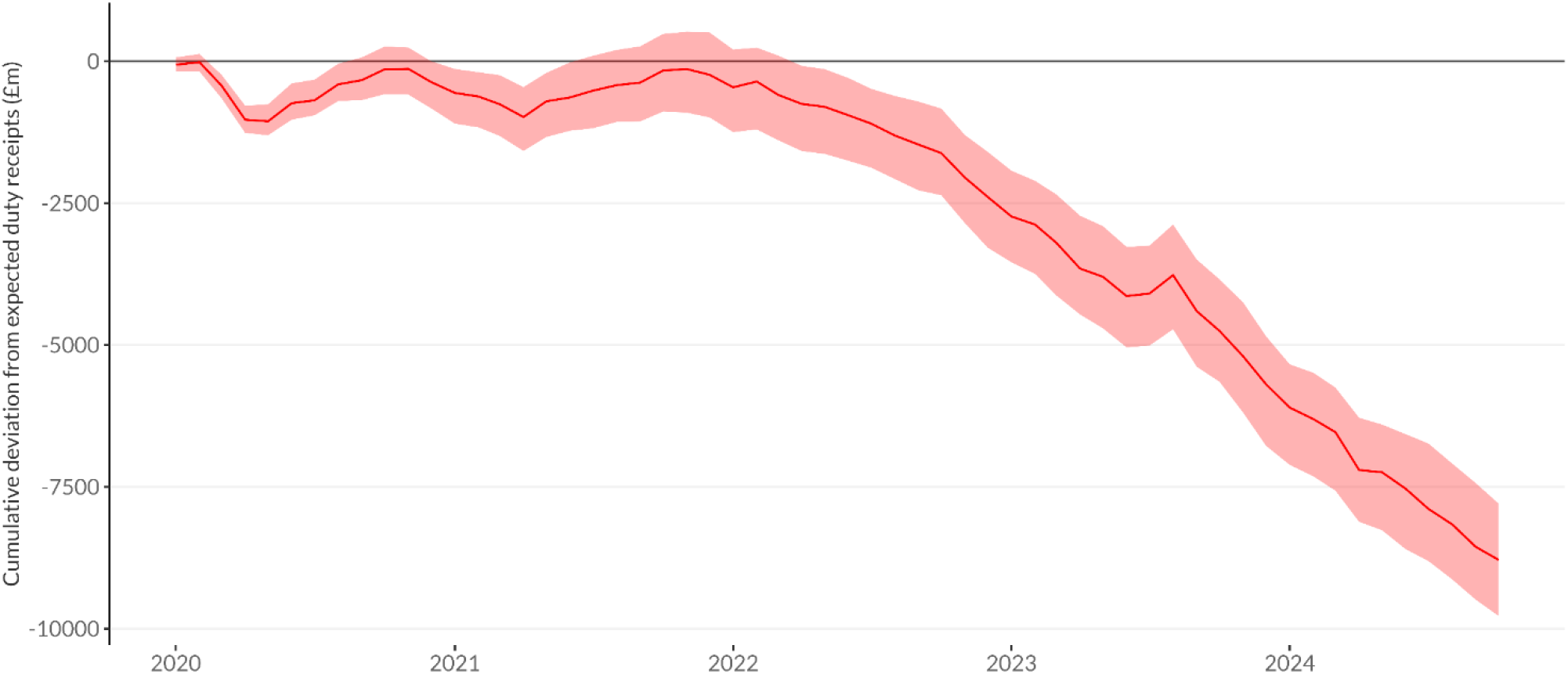
Cumulative excess duty revenue since January 2020 with 95% prediction interval.

Since the start of 2022 when inflation rates began to rise, duty receipts have consistently been below the levels that would have been expected based on pre-pandemic trends. This gap appears to widen in November 2022, when inflation rates peaked, and has remained at this higher level through into early 2024. The only deviation from this pattern was in August 2023, when duty receipts rose sharply. This almost certainly reflects alcohol producers choosing to pay duty earlier than they usually would on products stored in their warehouses in advance of the increases in alcohol duty that came into force that month, a process known as ‘forestalling’.

Figure 3 presents the monthly difference between observed and expected receipts since January 2020 by beverage type, with the equivalent cumulative difference shown in Figure 4. These show larger falls in duty receipts for beer during the COVID lockdowns, and larger increases in receipts for spirits in their immediate aftermath. Beer also shows comparatively smaller falls relative to the pre-pandemic trend since 2022 compared to wine and spirits. For wine, receipts have been consistently below expected levels since late 2021, with this deficit increasing gradually throughout 2022. In contrast, spirits receipts were at expected levels for most of 2022 until November, when a significant deficit appeared that has remained broadly stable ever since. For beers, wines and spirits, the fall in duty revenues stopped briefly in August 2023, with revenues in that month significantly higher than expected. This corresponds to the final month in which alcohol producers could clear products for sale (i.e. pay the duty on them) prior to the duty reforms and 10.1% increase in duty rates. However, Figure 4 illustrates that this represented a temporary outlier in the underlying trend rather than a shift in the direction of that trend.

**Figure 3.**
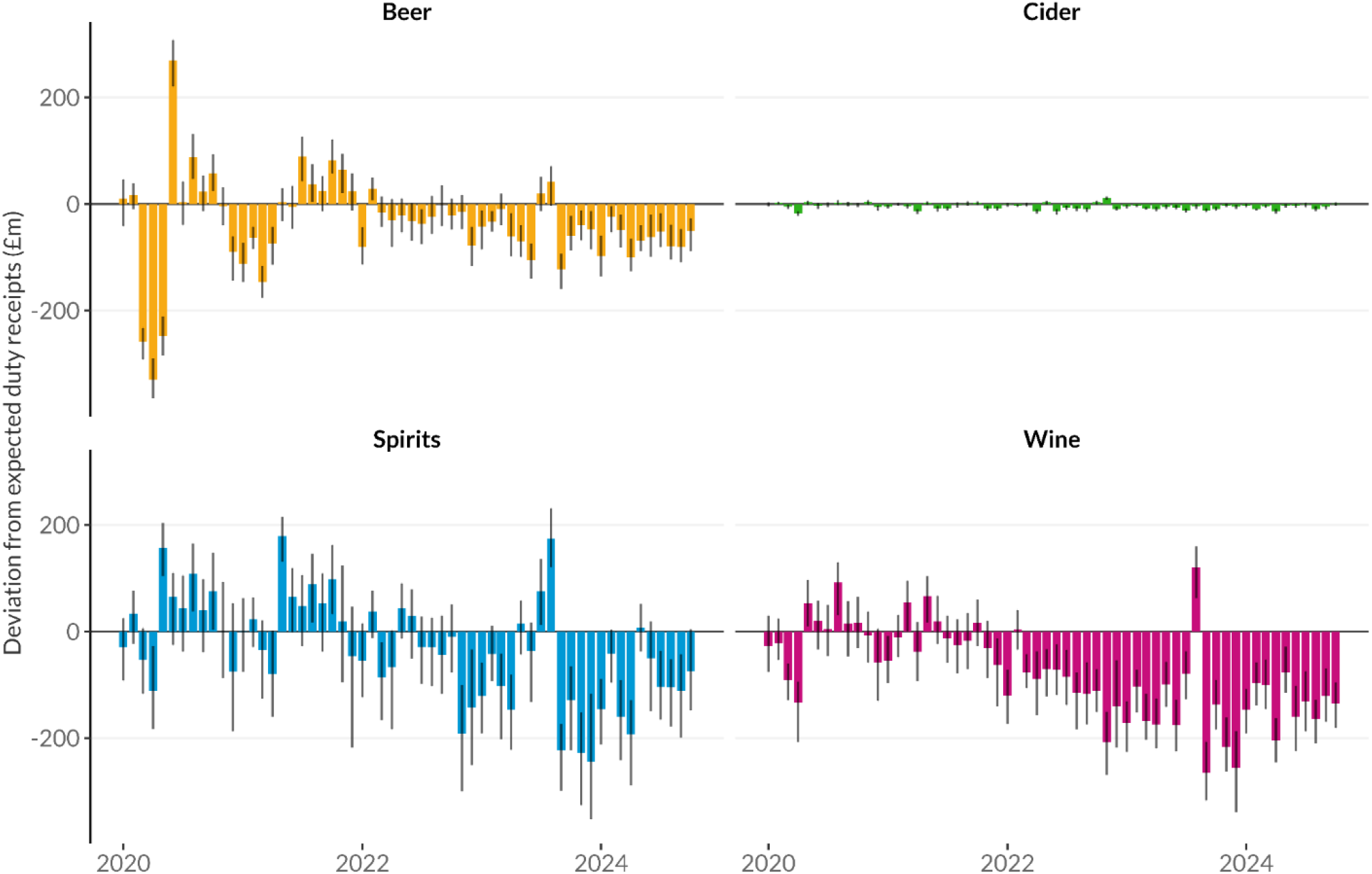
Excess monthly duty revenue in the UK by beverage type since January 2020 with 95% prediction intervals.

**Figure 4.**
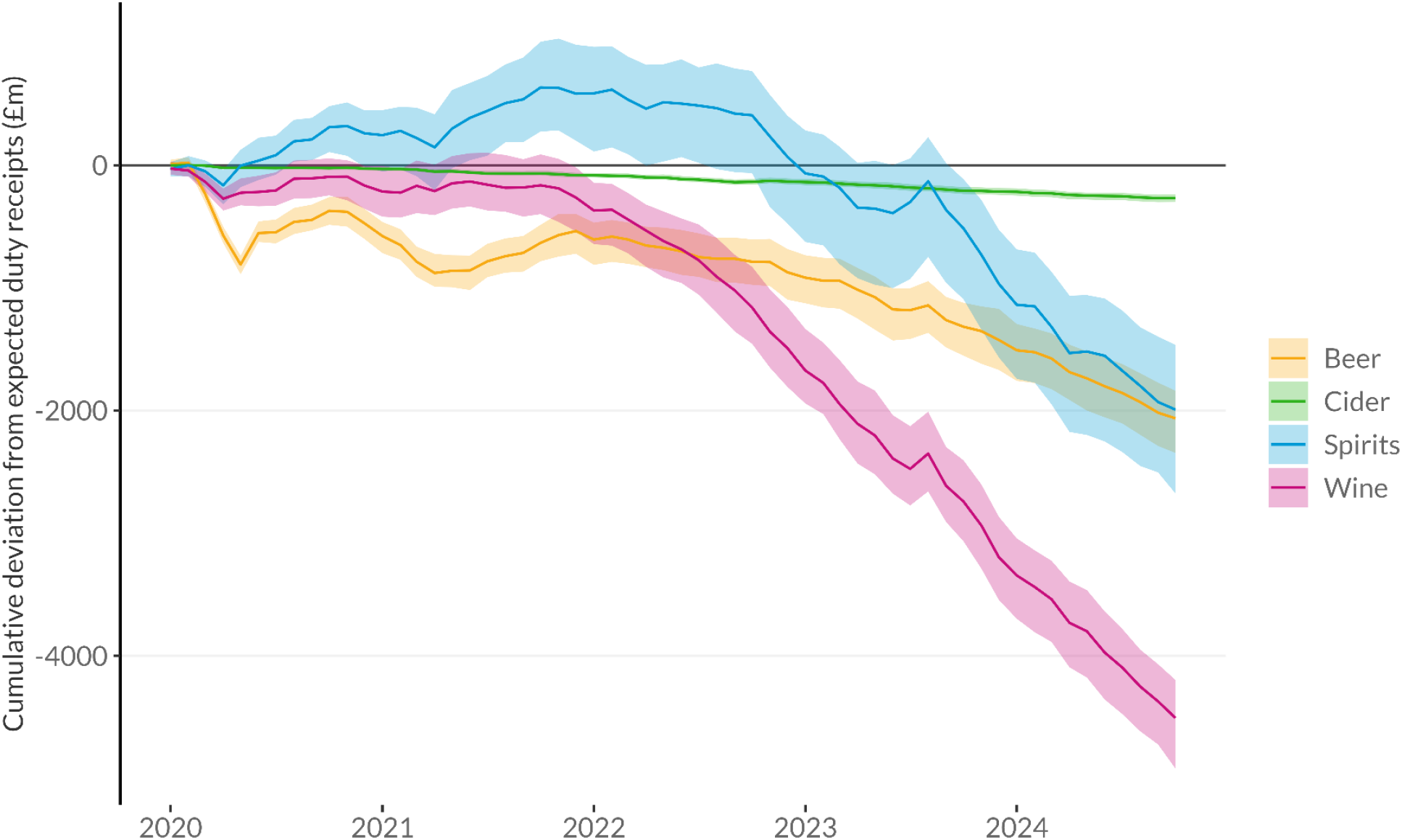
Cumulative beverage-specific excess duty revenue since January 2020 with 95% prediction intervals.

In a sensitivity analysis using only pre-2018 training data, to assess whether trends in receipts were already deviating from prior trends before the COVID pandemic, our results are broadly similar, with no clear indication of a change in overall duty revenue trends prior to 2020. Beverage-specific results suggest that beer receipts may have been rising above pre-2018 trends and wine receipts below pre-2018 trends before any impact of the pandemic, however the changes in trends during the pandemic and the cost-of-living crisis identified in our primary analysis remain evident.

## Discussion

Our analysis shows that, relative to pre-pandemic trends, government revenue from alcohol duties was relatively unchanged overall across 2020 and 2021, although revenues fell during periods of lockdown with corresponding increases as restrictions were lifted. The falls in revenue were greatest for beer, most likely due to the fact that pubs were closed during lockdowns and beer is consumed disproportionately in the on-trade^15^. Meanwhile spirits revenues increased above expected levels over this period, with the largest rises in period when restrictions were less stringent.

More recently, overall alcohol duty revenues since early 2022 have fallen relative to the pre-pandemic trend, with a cumulative gap of around £8.8 billion overall. This suggests that alcohol sales are likely to have fallen, on average, during the cost-of-living crisis. Previous studies have shown two key mechanisms at play in relation to alcohol consumption during economic downturns: reductions in consumption as individuals cut back on non-essential spending in the face of financial pressure and increases in consumption in response to increased stress driven by economic challenges^16^. Recent data suggests 7 million adults in Great Britain (one in eight) reported experiencing financial hardship during 2022 and/or early 2023 and that this was strongly associated with higher rates of psychological distress^17^. Our analysis suggests a greater degree of cutting back than drinking to cope with stress, at least at the population level, although there may still be important differences between individuals. The potential for this is highlighted by the fact that by far the largest cumulative reduction in duty revenues compared to expectation is in wine, which is more commonly drunk by those in higher socioeconomic groups^18,19^. This is somewhat contrary to expectations, as it is lower socioeconomic groups who have been most affected by the cost-of-living crisis^7^. It is also surprising that wine receipts have fallen gradually below expected levels since late 2021, while spirits, and to a lesser extent beer, receipts only fell below expected levels in November 2022, suggesting that there may be different underlying causes for these trends. Further research is needed to examine individual changes in alcohol consumption over this period to understand the impact of these economic challenges and the resulting implications for alcohol harms.

Finally, our results show clear evidence of ‘forestalling’ or bringing forwards of the payment of alcohol duties by the alcohol industry in relation to the August 2023 duty reforms, with large volumes of alcohol apparently being cleared for sale immediately prior to the reforms that would likely otherwise have been cleared in the months thereafter. However, there is no clear evidence of a longer-term impact of the reforms on duty receipts, rather than a continuation of pre-reform trends, contrary to claims made by the alcohol industry^20^.

Our analysis used robust data and a novel approach to examine trends in alcohol duty receipts up to October 2024, however there are several important limitations. Most importantly, although there is a strong correlation between duty revenues and alcohol sales volumes, receipts are not a direct measure of alcohol sales or consumption. Although spirits and beer are taxed directly on the basis of alcohol content, wine and cider duty prior to the duty reforms was based on product volume. As a result, if consumers had shifted from higher- to lower-strength wines, or vice versa, this would not be captured in duty receipts. Further, these figures relate to alcohol cleared for sale, not alcohol sold and therefore do not account for potential stockpiling by either alcohol producers/retailers or by consumers. It is also possible that differential price changes between beverage types may have driven shifts in sales between products, although beverage-specific inflation indices published by the UK Government do not suggest large deviations in prices over time by beverage (see Figure A4).

Overall our findings suggest little overall change in alcohol sales in the UK during the COVID- 19 pandemic relative to pre-pandemic trends, but a clear fall in sales during the cost-of-living crisis driven initially by wine and, more recently, spirits. We find little evidence that the duty reforms have impacted on pre-reform trends.

## Data Availability

All data is publicly available. R code to replicate our analysis is available at https://github.com/VictimOfMaths/Experiments/blob/master/HMRCReceiptsAnalysis.R

## Funding

This work was supported by the National Institute for Health and Care Research (NIHR156679). The views expressed are those of the author and not necessarily those of the NIHR or the Department of Health and Social Care.

## Conflicts of Interest

None to declare

## Online Supplementary Material

**Figure A1.**
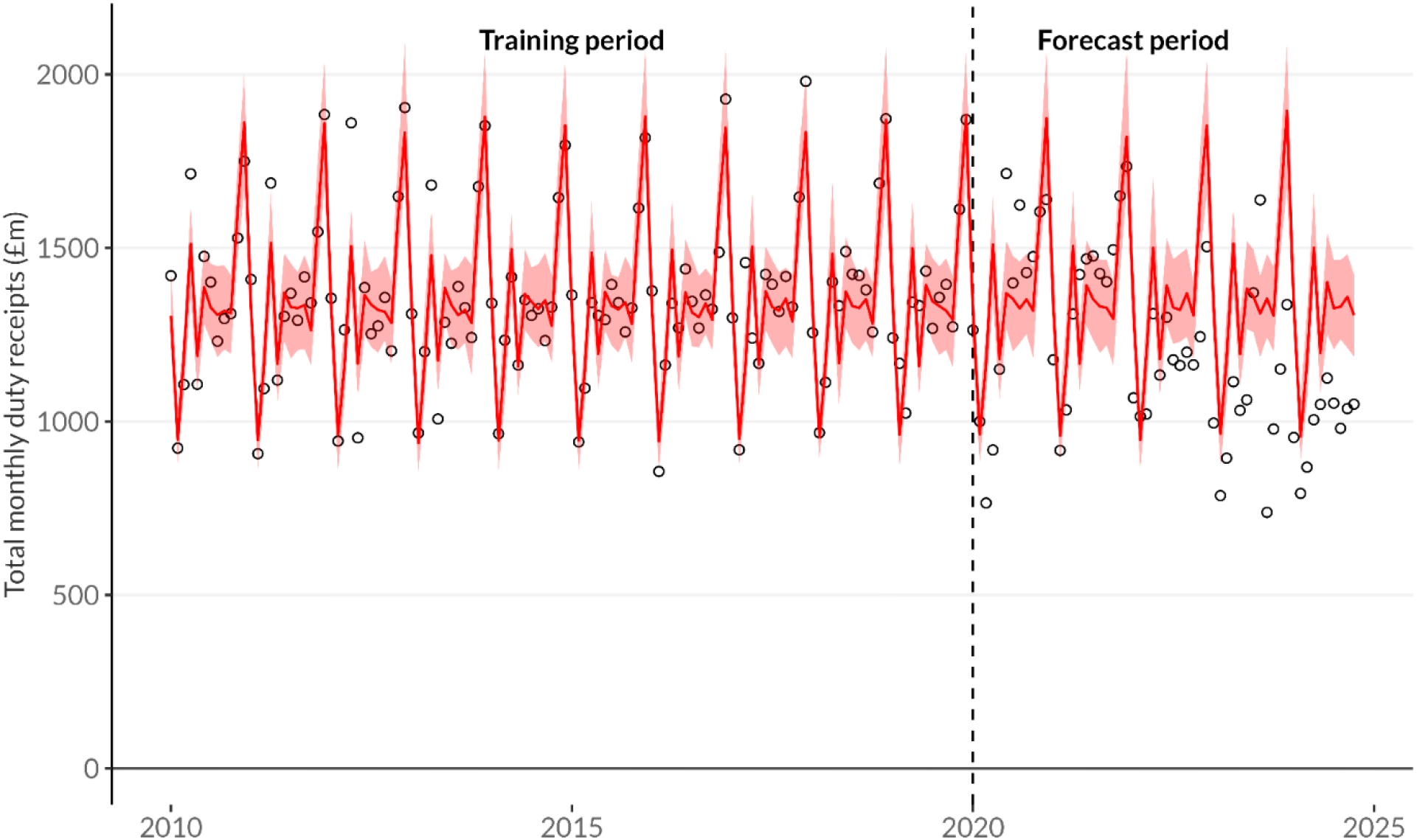
Modelled (red line with 95% prediction intervals) and expected (black circles) alcohol duty receipts. Vertical dashed line represents the end of the training data period.

**Figure A2.**
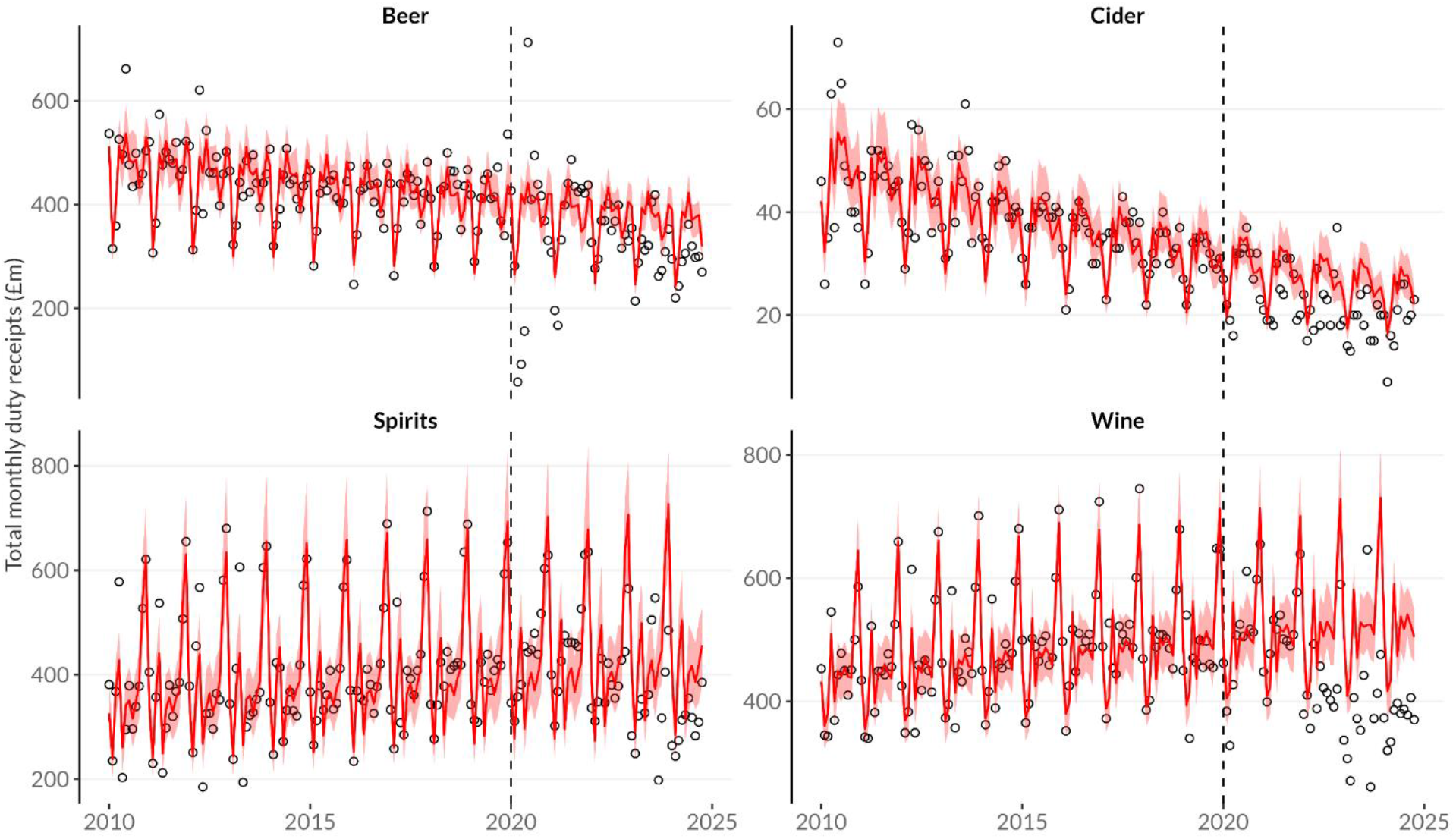
Modelled (red line with 95% prediction intervals) and expected (black circles) alcohol duty receipts by beverage type. Vertical dashed line represents the end of the training data period.

**Figure A3.**
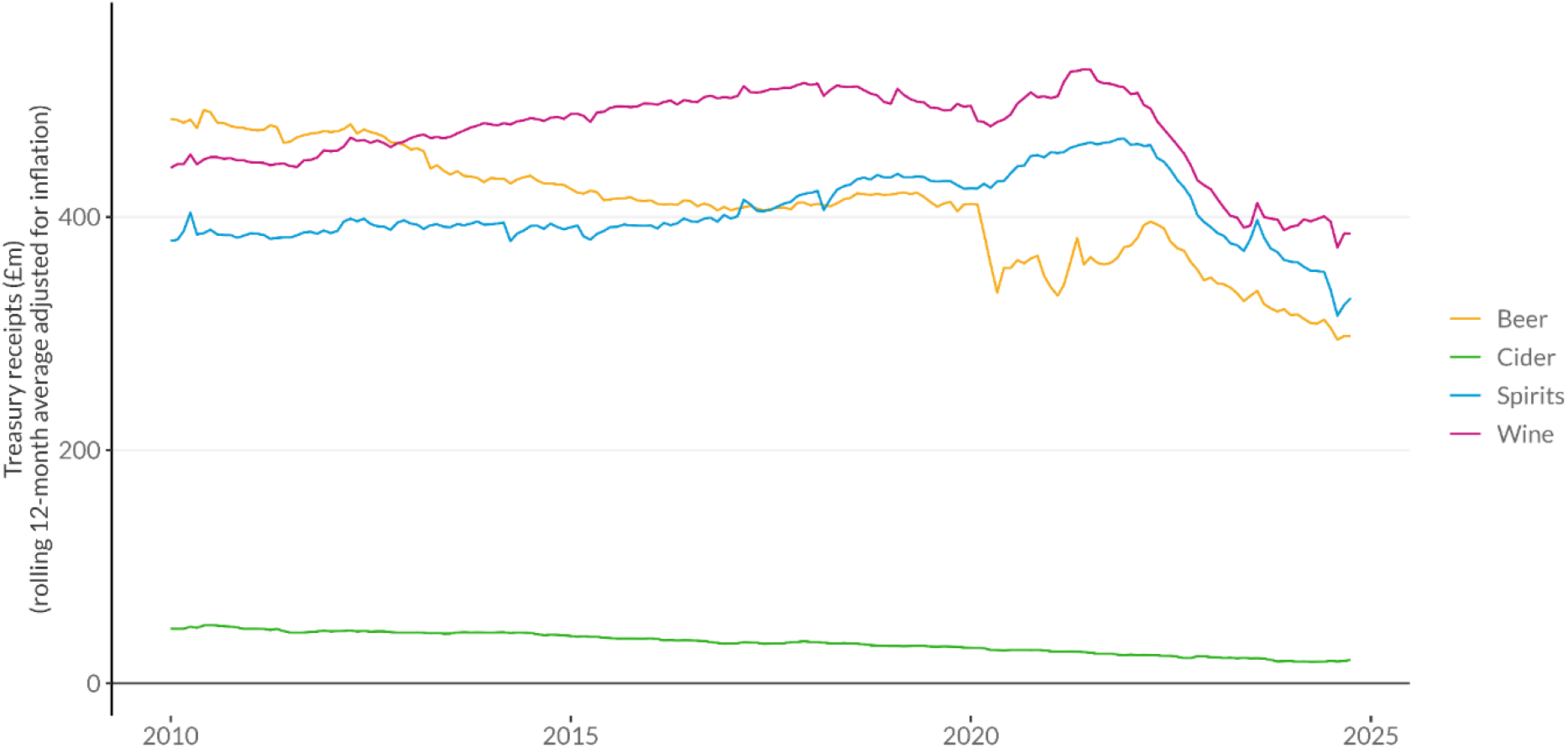
Raw alcohol duty receipts (12-month rolling average) 2000-2024.

**Figure A4.**
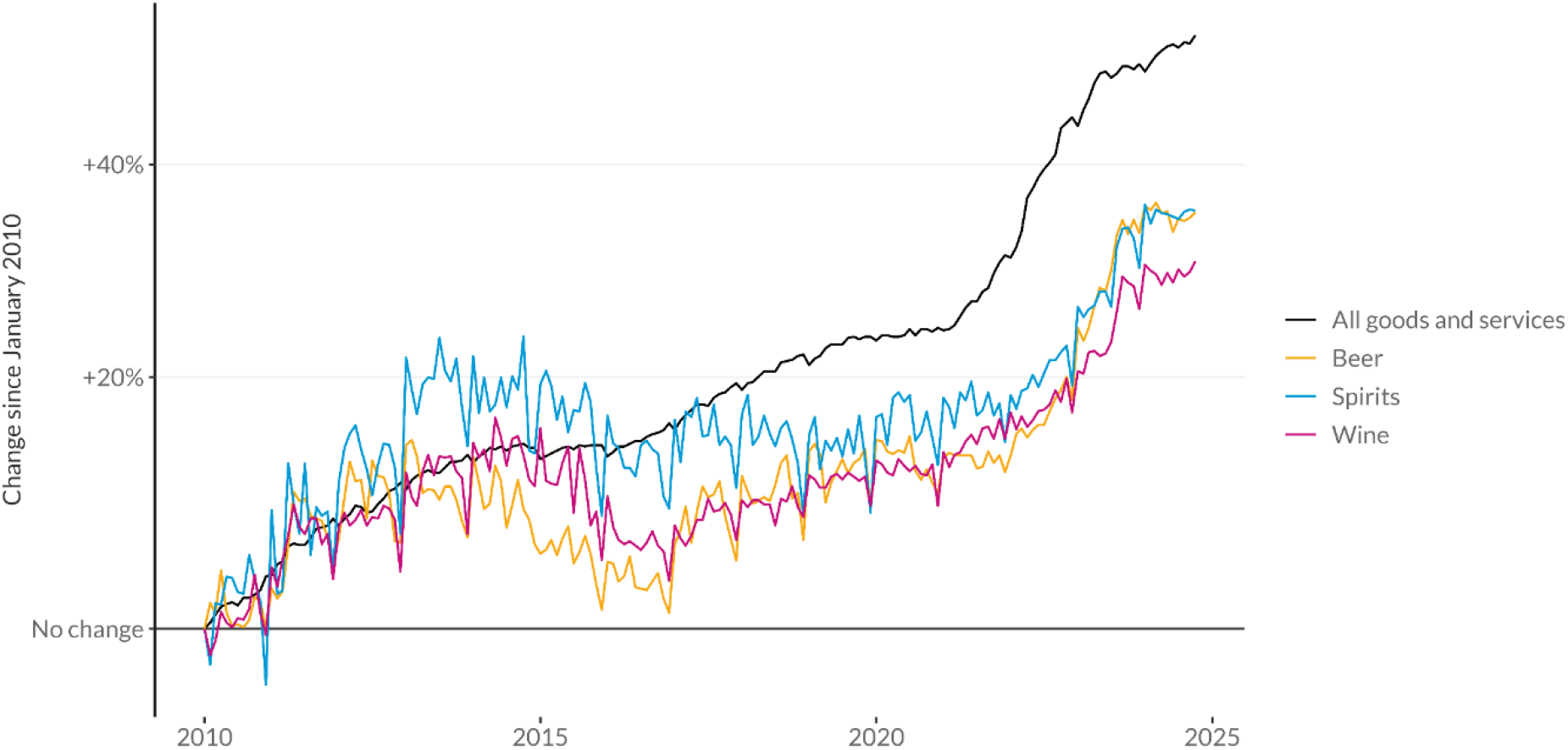
Changes in CPI inflation since January 2010 for alcohol and all goods and services.

### Sensitivity analysis using training data up to December 2017 only

**Figure A5.**
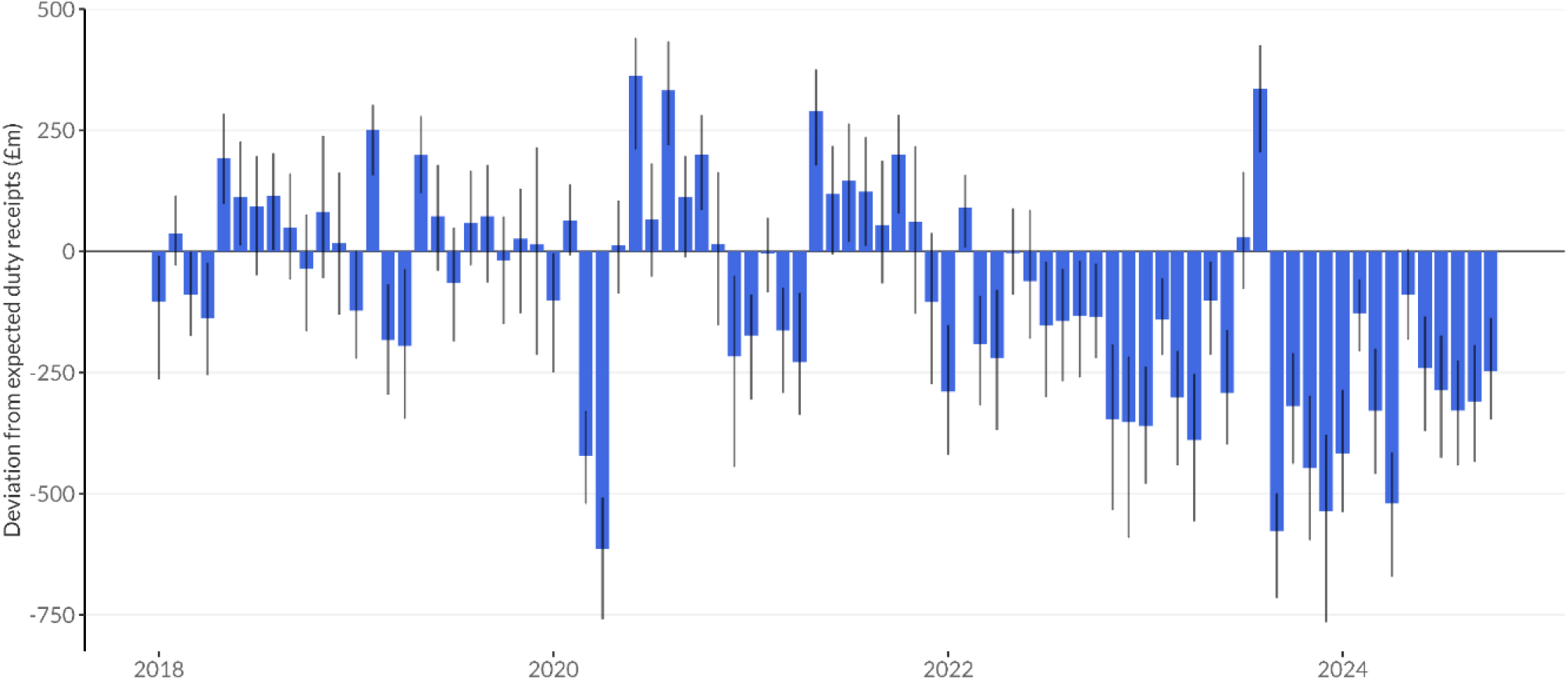
Excess monthly duty revenue in the UK since January 2018 with 95% prediction intervals.

**Figure A6.**
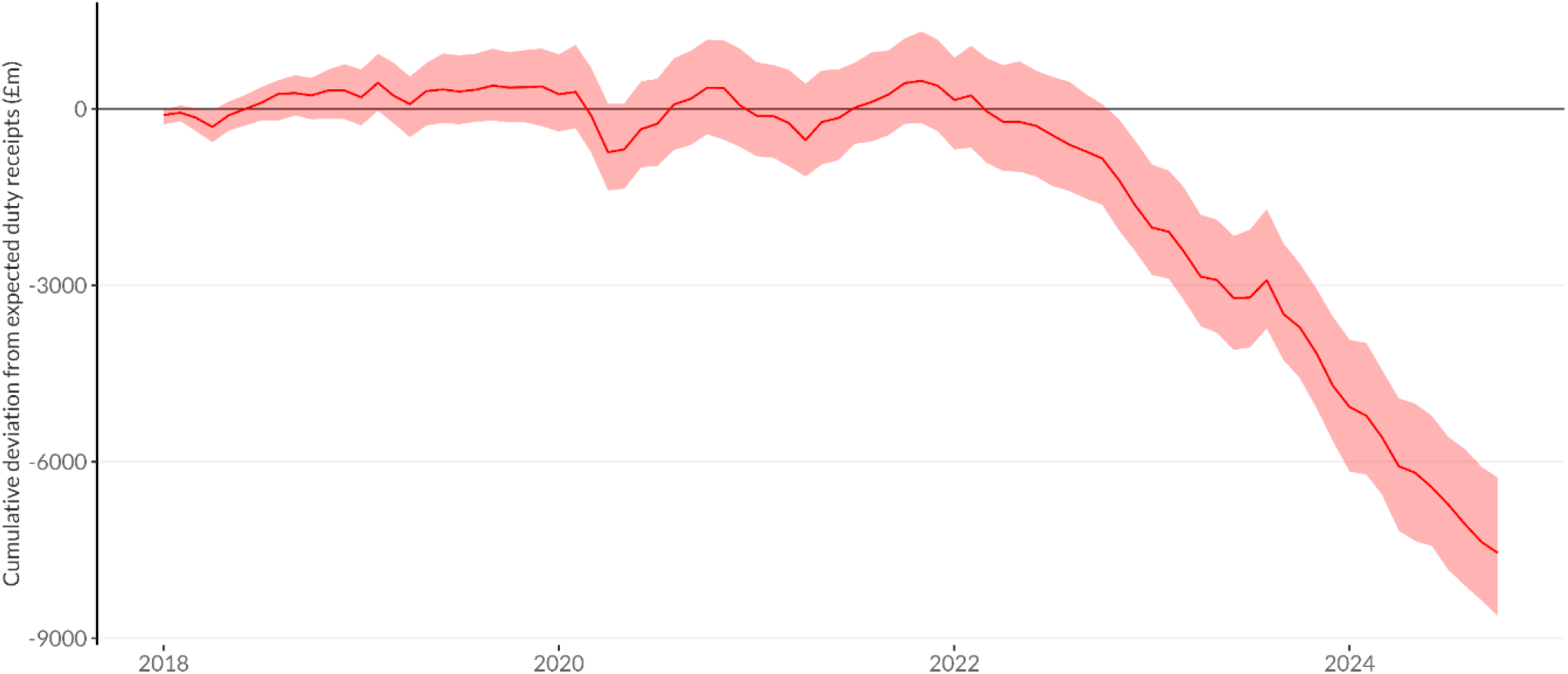
Cumulative excess duty revenue since January 2018 with 95% prediction interval.

**Figure A7.**
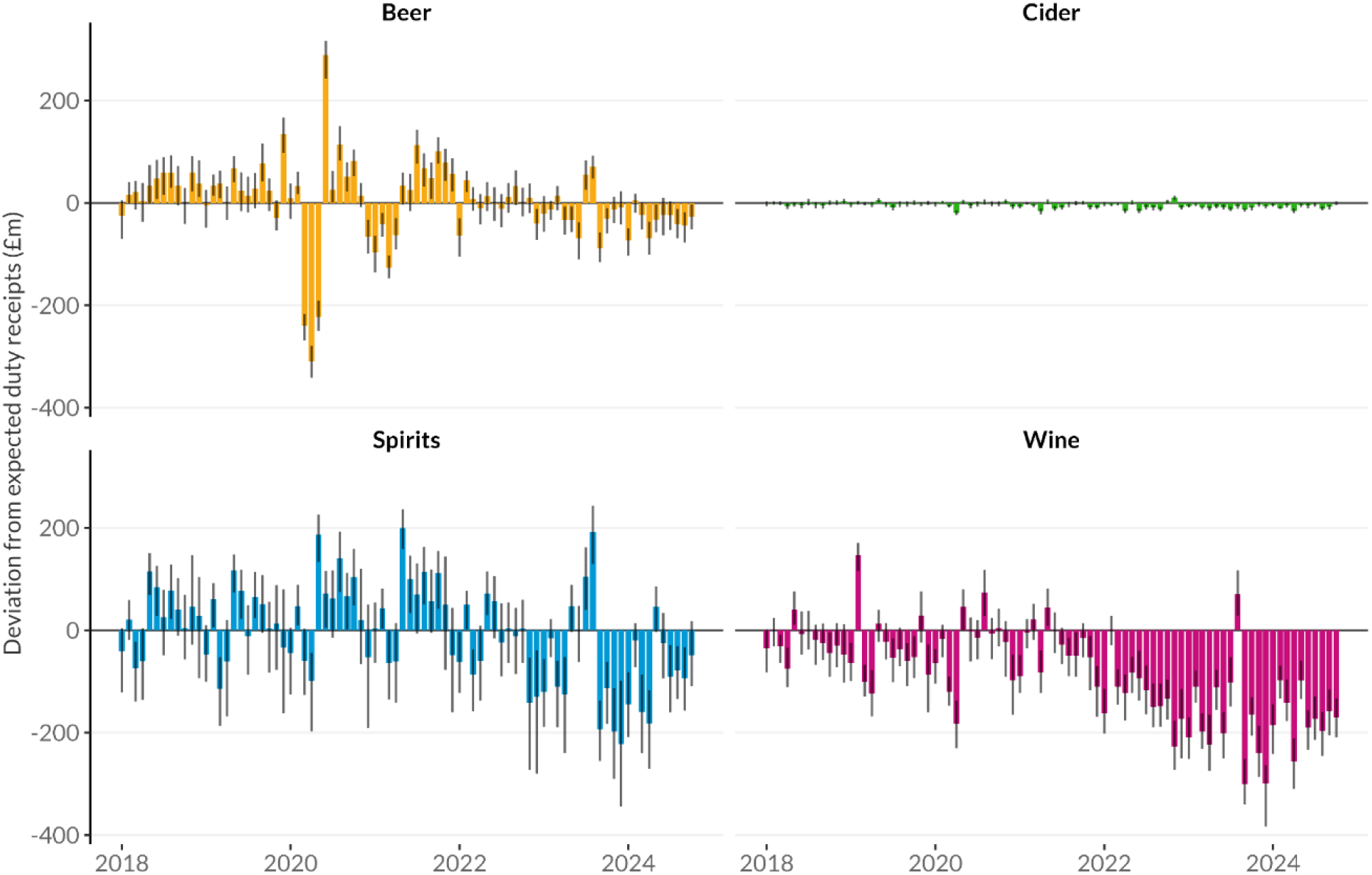
Excess monthly duty revenue in the UK by beverage type since January 2018 with 95% prediction intervals.

**Figure A8.**
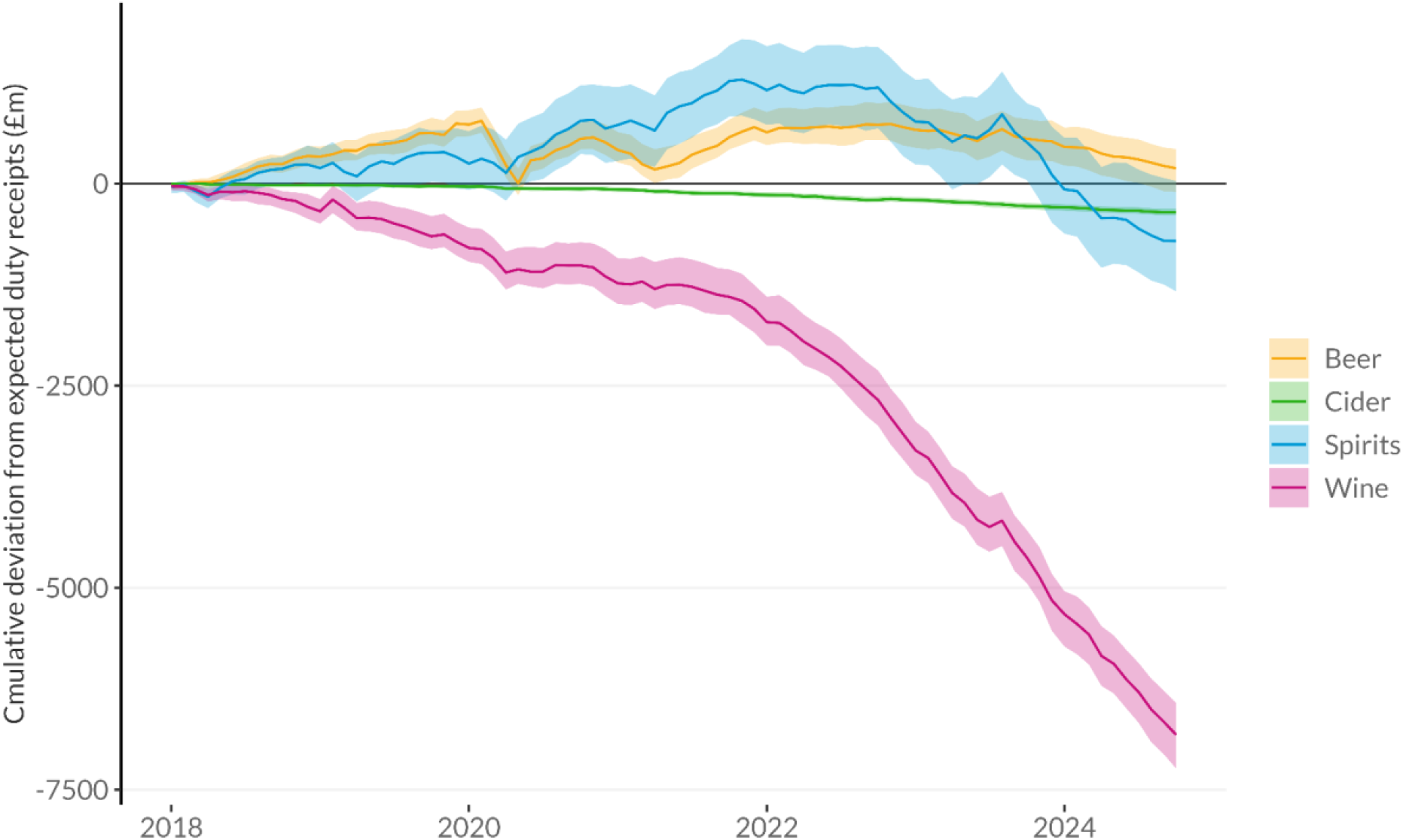
Cumulative beverage-specific excess duty revenue since January 2018 with 95% prediction intervals.

## Notes

### Competing Interest Statement

The authors have declared no competing interest.

